# Prevalence, perceptions and factors influencing covid-19 vaccines’ uptake among nurses in fako division, cameroon

**DOI:** 10.1101/2023.01.25.23284999

**Authors:** Niba Clinton Ambe, Achidi Eric Akum, Nkemayim Florence Binwi, Palle John Ngunde

**Affiliations:** Department of Nursing, Faculty of Health Sciences, University of Buea, South West Region, Cameroon; Department of Biochemistry and Molecular Biology, Faculty of Science, University of Buea, South West Region, Cameroon; Department of Nursing and Midwifery, Faculty of Healthcare Professions, H.O.P.E University Institute (HUI) Limbe, South West Region, Cameroon

**Author notes:** **Corresponding Author:** (NCA).

**Keywords:** Nurses, Perceptions, Prevalence, Factors, COVID-19 Vaccines Uptake, Fako Division

## Abstract

**Background:** Since December 2019, the world has watched the rapid growth of a new pandemic, the COVID-19, a novel pandemic causing death and disruption of normal life. This COVID-19 continues to spread and poses serious threats to public health throughout the world. Even though vaccines are available, there is no guarantee of critical population vaccination, as there still exist stiff resistance to the uptake of the currently available vaccines.

**Purpose:** To assess nurses’ perceptions regarding the uptake of COVID-19 vaccines, determine the proportion of nurses vaccinated, as well as the associated factors influencing the uptake of COVID-19 vaccines, so as to alert decision makers on the possible limitations associated with the uptake of the vaccines in the nursing population in Fako Division, Cameroon.

**Methods:** This was a mixed method quantitative and qualitative study conducted in Fako Division. A multistage random sampling technique was employed to enroll participants into the study. We collected quantitative data from consented nurses through the use of a structured questionnaire from April 06^th^ to June 2^nd^, 2022, and qualitative data from nurse leaders through focused-group discussion from June 3^rd^ to 23^rd^, 2022. In the quantitative phase, we performed descriptive and inferential statistics using the SPSS Version 23.0 and in the qualitative phase, we performed a thematic content analyses and transcription.

**Results:** In the quantitative phase, we had more females 131(66.5%), and nurses aged 26-40years 90(45.7%). Most nurses worked in the maternity 49(24.9%). In the qualitative phase, 20(76.9%) were females. Regarding nurses perceptions of COVID-19 vaccines uptake, 133(67.5%) nurses had positive perceptions, and 26(07.6%) had “No trust” in the vaccines. Most nurses 109(55.3%) had not been vaccinated. Negative factors reported by nurses included the belief that the vaccines were dangerous and could cause death 120(60.9%) and 32(16.2%) said no one can influence them to change their minds about taking the vaccines. In the qualitative phase, a majority of the nurse leaders 15(57.7%) wished that COVID-19 vaccination should continue, but with accompanying research to eliminate side effects. It was observed that a majority of the nurse leaders 16(61.5%) had been vaccinated. The respondents reported some factors (belief factors, social influence and lack of knowledge), which had negatively influenced them from taking the COVID-19 vaccines.

**Conclusion:** Nurses perceived high relevance for the COVID-19 vaccines while a majority of the nurse leaders perceived that the COVID-19 vaccines are not safe, ineffective with numerous side effects, has a magnetic effect, politically motivated with bad faith, and has the possibility to cause infections. Furthermore, most nurses had not been vaccinated, but a majority of nurse leaders were reported to have taken a COVID-19 vaccine. Several negative factors including belief, social influence and religious factors were reported to have contributed to the lower uptake of the COVID-19 vaccines amongst nurses and nurse leaders in Fako division, Cameroon.

## Background

Since December 2019, the world has watched the rapid growth of a new pandemic disease, coronavirus disease 2019 (covid-19), a novel pandemic causing tremendous suffering, death, and disruption of normal life [1]. The COVID-19 viral disease is amongst the deadliest infectious diseases to have emerged in recent history. As with all past pandemics, the specific mechanisms of its emergence in humans remains unknown [1]. This virus belongs to the B-coronavirus group, the same group of the 2003 Severe Acute Respiratory Syndrome Coronavirus (SARS-CoV), and for the similarity, it was named SARS-CoV-2 [2]. Genomic analysis suggests that, the COVID-19 virus originated in bats and was transmitted to humans through an unknown intermediate hosts in Wuhan, China.

According to WHO [3], the COVID-19 disease affects different people in different ways. Most infected people will develop mild to moderate illness and recover without hospitalization. However, serious symptoms such as breathing difficulty or shortness of breath, loss of speech or mobility, confusion and chest pain posed serious risk to the patient. Since its discovery, more than 2,850,000 cases have been reported, including nearly 200,000 deaths [4, 5]

Globally, the covid-19 pandemic, caused by SARS-CoV-2 continues to spread and poses serious threats to public health and economic stability throughout the world. Thus, to protect the global population, developing safe, effective and acceptable vaccines is mandatory to control the spread of SARS-CoV-2 pandemic [6]. The development of vaccines is considered among the greatest scientific achievements in promoting global health [7], with vaccination programs decreasing the burden of infectious diseases, loss of life, and contributing to strengthening the health system as a whole. However, reaping such benefits depends upon a willingness of the population to be vaccinated [8]. Even as vaccines are available, Omer *et al*., [9] realized that the availability of vaccines does not guarantee sufficient population vaccination as evidenced by vaccine hesitancy which stems from multiple key factors including; complacency (the person does not see a need and value for the vaccine), individual’s lack of confidence in the vaccine, as well as convenience (access to vaccines) [10]. More so, previous research had proven that vaccine uptake remains variable and inconsistent, and successful inoculation against this disease will require widespread public educational campaigns regarding vaccine safety and efficacy [11].

Dror [11] in his 2020 study argued that, people who were currently vaccinated against seasonal influenza had a strong tendency to accept a future COVID-19 vaccine, but Healthcare Worker’s (HCWs’) perceptions of COVID-19 vaccines uptake in his study was intriguing. The rate of acceptance for a COVID-19 vaccine among physicians and nurses overall was lower than their acceptance rates of seasonal influenza vaccination. Other studies regret that voluntary vaccination had been sub-optimal among HCWs in the United States, a priority group for whom immunization is essential for maintaining health system capacity and the safety of high-risk patients in their care [12].

Africa is no exception. In the context of the Democratic Republic of Congo, a study of 613 HCWs, including 50.9% men and 49.1% women revealed that, only 27.7% of the HCWs said that they would accept a COVID-19 vaccine if it was available. In this entire population, up to 72.8% of the HCWs were nurses which was a significant predictor to consider [13]. Considering other schorlarly works conducted in Nigeria, many HCW workers demonstrated unwillingness for the COVID-19 vaccines; a major setback to attain full immunization coverage among HCWs. However, knowledgeable HCWs with high motivation and high willingness reported a 15-times higher odds ratio of being fully vaccinated and by comparison, motivation relatively contributed to their high uptake of COVID-19 vaccines. Much of the recent literature coming out of Nigeria and other low-middle-income countries (LMICs) focused on increasing motivation as a factor to get a COVID-19 vaccination [10].

In a Cameroonian study by Dinga *et al*., [14], vaccine hesitancy to a COVID-19 vaccine was 84.6% of 2512 respondents. Reasons for vaccine hesitancy using the WHO matrix of vaccines hesitancy determinants included communication and media environment, perception of pharmaceutical industry, reliability and/or source of vaccine and cost. Most Cameroonians raised concerns of safety, efficacy and confidence. Among 371 healthcare workers who took part in another study, only 45.38% indicated willingness to accept the vaccine if offered [15].

However, eradicating the COVID-19 disease and ensuring patients safety in the hands of the HCWs, will require a safe vaccine which is first acceptable by nurses since they are the frontline care provider. In doing so, it is imperative to appraise the proportion of nurses and nurse leaders vaccinated with a COVID-19 vaccine, their perceptions about the vaccines and factors associated with COVID-19 vaccines’ uptake in the context of Fako Division, Cameroon.

## Materials and Methods

### Study design

In this study, a Mixed Method Research (MMR) of qualitative and quantitative design was employed, from october 2021 through July 2022. A cross-sectional survey was used in the quantitative research design to describe the overall picture of a phenomenon at a given point in time, and data was collected with the use of a structured questionnaire. On the otherhand, focused-group discussion was employed for the qualitative aspect of the research.

### Study population

The sample population included all nurses (quantitative phase) and nurse leaders (qualitative phase), working in the Tiko District Hospital (TDH), Buea Regional Hospital (BRH) and the Limbe Regional Hospital (LRH) in Fako Division, Cameroon.

### Selection criteria and sample size calculation

For the qualitative research, we included nurse leaders (unit heads or ward charges or assistants, and general supervisors or assistants) in the selected health facilities. These persons are charged with the general supervision of the hospital, management of units and the coordination of COVID-19 vaccination program. Nurse leaders who consented and were available to participate in the study were enrolled. For the quantitative phase, ward nurses working in the selected facilities that were available during data collection and consented to take part in the study were recruited. However, we excluded nurses and nurse leaders in the selected facilities who gave their consent to be part of the study but were sick of COVID-19 or other major bed-bound illnesses at the time of data collection.

A sample size of 203 nurses for the quantitative phase was calculated based on the formula proposed by Cochran (1963:75) [16]. However, the finite sample size calculation was done for the various selected facilities in the different health district in Fako Division, using the probability proportionate to sample size sampling method (Tab 1). This produced separate sample sizes for the different health facilities.

A total of 27 nurse leaders were recruited for the qualitative study in the three hospital facilities. Three separate Focused-Group Discussions (FGDs) was conducted for the three different hospitals. Each FGDs had one (1) or two (2) general supervisors, seven (7) Unit Heads, and one (1) coordinator in charge of COVID-19 vaccination. General supervisors in each facility served as moderators during the discussion.

**Tab 1:** Sample size calculation for the study (probability proportionate to size)

| SN | Hospital Category |  |  | Proportional Sample size<br>for the quantitative study<br>(For each Facility) |
| --- | --- | --- | --- | --- |
|  |  | Actual Number of Registered<br>Nurses in the Selected<br>Facilities. | Proportion |  |
| 1 | Limbe Regional Hospital | 214 | 0.50 | 102 |
| 2 | Buea Regional Hospital | 180 | 0.42 | 85 |
| 3 | Tiko District Hospital | 34 | 0.08 | 16 |
| Total Actual Population |  | n= 428 | 1 | n= 203 |

### Ethical Considerations

Ethical clearance was obtained from the Faculty of Health Sciences, University of Buea Institutional Review Board (UB-IRB) with IRB number 2022/1833-05/UB/SG/IRB/FHS for the protection of human rights. Administrative authorization was gotten from the Regional Delegation of Public Health for the South West Region and the Regional Directors, and departmental heads of the Hospitals where the study was carried. Nurses were adequately sensitized on the objectives and protocol of the study. Those who consented were then enrolled into the study. All information in this study was given code numbers and were anonymously recorded.

### Data processing and analysis

The data collected was thoroughly checked for accuracy, completeness, distribution, validity and reliability of measures before analysis. For the qualitative phase, Thematic Content Analysis (TCA), transcription and data cleaning was performed. Findings were categorized in themes, subthemes, grounded and then summarized in quotations. For the quantitative phase, data screening and cleaning was completed by exposing the data and later entered into SPSS Version 23.0 for analysis. Descriptive analysis involved tables, graphs, charts, percentages, and frequencies to indicate distribution of variables and mean for continuous variables like age. Significance of p-values was designated at P<0.05.

## Results

### Socio-demographic data

In the quantitative study, we had more females 131(66.5%). A majority of the nurses 90(45.7%) were aged 26-40years, followed by those 18-25years 67(34.0%). Single nurses constituted the majority 101(51.3%), followed by those married 67(34.0%). Most nurses were working in the maternity 49(24.9%), medical 33(16.8%) and surgical 30 (15.2%) units and a large proportion 89(45.2%) of the nurses had worked for between 5-10years

In the qualitative phase, females 20(79.92%) were by far more than males 06(23.08%). A majority of the nurses 18(69.23%) were aged 26-40years, followed by those aged >50years 05(23.08%). Regarding their level of training, a majority held a bachelor’s degree 12(46.15%) and most of our respondents were working at the COVID-19 vaccination/maternity/ANC units 07(26.93%). The respondents were of the Christian background and distributed by health facilities as follows: TDH 08(30.77%), BRH 09(34.62%) and the LRH 09(34.62%)

### Nurses’ perceptions regarding uptake of COVID-19 vaccines

Regarding nurses’ thoughts/perceptions on the uptake of the COVID-19 vaccines; 133 (67.51%) of the nurses had positive perception; 74(37.5%) were confident they will take the vaccines when exposed to infected cases, 25(16.8%) indicated that they will take it if they understood its benefits and just 15(07.6%) will take the vaccines if the disease had proved severe on others. Some said they were just so overwhelmingly confident 19(05.6%) about taking the vaccines. However, a cumulative percentage of 57 (28.93%) had negative perceptions about the vaccines, with most nurses reporting “No trust in the vaccines (ingredients & pharmaceutical companies)” 26(07.6%) (Tab 4).

**Table 2:** Nurses perceptions regarding uptake of COVID-19 vaccines in Fako Division.

| SN | Indicators | Responds | Frequency n (%) |
| --- | --- | --- | --- |
| 1 | The need for nurses in Cameroon to take the vaccines | Yes | 111 (56.3) |
|  |  | No | 86 (43.7) |
| 2 | Nurses thoughts/ perceptions in taking COVID -19 vaccines | Will take when exposed to Infected cases. | 74(37.5) |
|  |  | Will take once I see the disease proves serious. | 15 (7.6) |
|  |  | Once I fully understand the benefits to me or general, I will take. | 25 (16.8) |
|  |  | Overwhelmingly confident about the vaccine. | 19 (5.6) |
|  |  | No trust (ingredients & pharmaceutical companies). | 26 (7.6) |
|  |  | Not safe and effective. | 11 (12.7) |
|  |  | The vaccines have dangerous side effects. | 13 (13.2) |
|  |  | Can instead cause infection to me | 7 (6.6) |
| 3 | Possibility of a nurse taking a COVID-19 vaccine. | Yes | 96 (48.7) |
|  |  | No | 101 (51.3) |

### Nurse leaders’ perceptions regarding uptake of COVID-19 vaccines

There are still negative perceptions regarding the uptake of COVID-19 vaccines amongst nurse leaders. *“We hope for the vaccination program to stop. It is not helping and has no Vaccine’s Vail Monitor (VVM)*” (06respondents).

*They reported and I quote “Why should I take vaccines without VVM: How do we know if it’s not a placebo?” (2Unit heads). “I can’t really say, vaccination should stop, but I think it is not helping for now” (GS, 1 Unit Head)*. Other perceived that the vaccines are politically motivated (06 respondents). *“This thing is political. Where have you seen or ever heard that people are paid to take vaccines? (3unit heads). “We Africans just follow the whites sometimes even in what we don’t understand. I perceive a political benefit from some people” (1unit head)*.

However, some said the vaccines were ineffective, had hash side effects, and could cause COVID-19 and even death (13respodents). There was a high perceived threat concerning the vaccines. *“We have managed patients who were vaccinated and the side effects kept some bedridden” (6unit heads) and one even died (2 unit heads). “Even when I took the vaccines, I felt like I was going to die with pains and the worst is that I still got infected thereafter” (3unit heads)* (Tab 8).

**Table 3:**
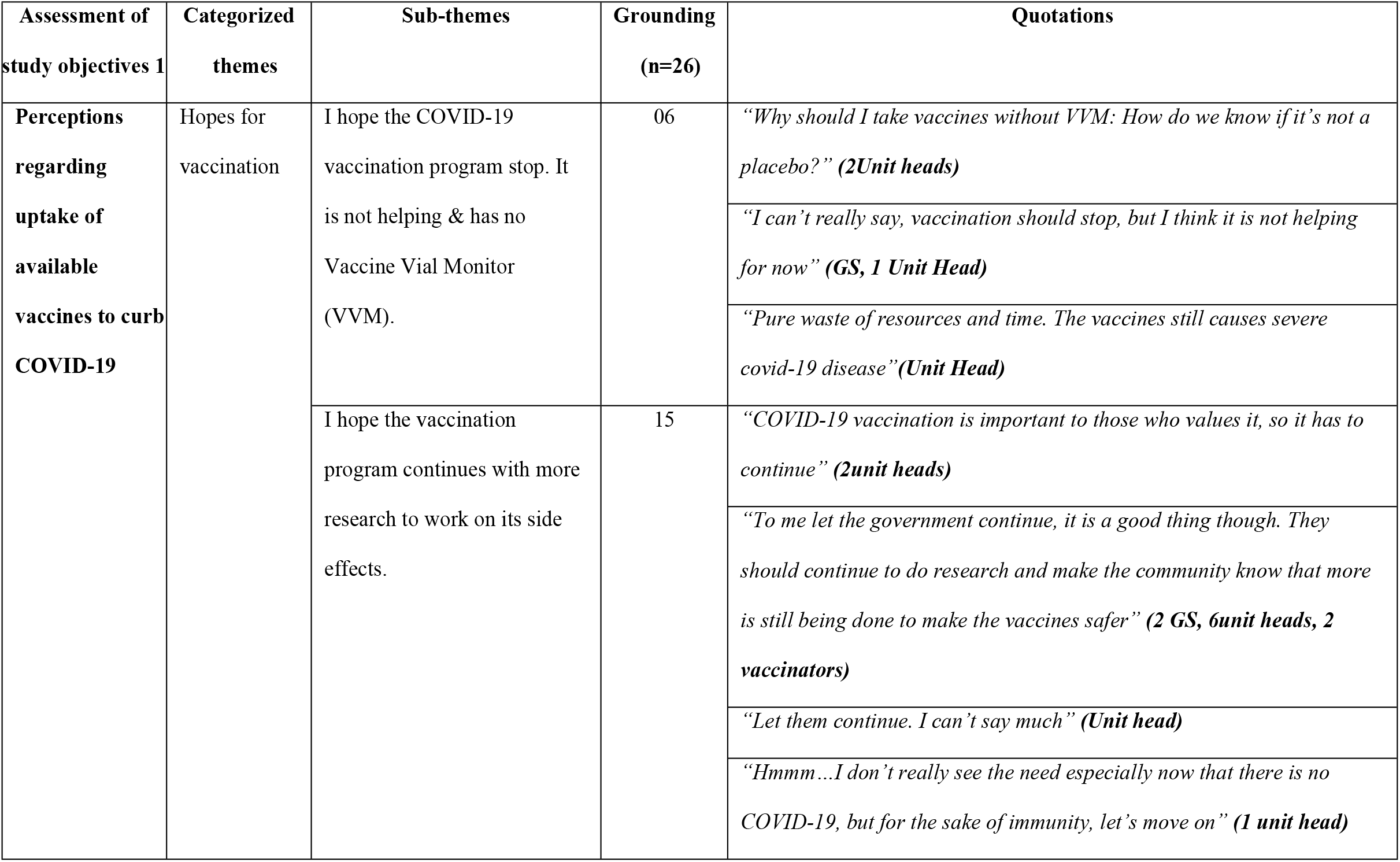
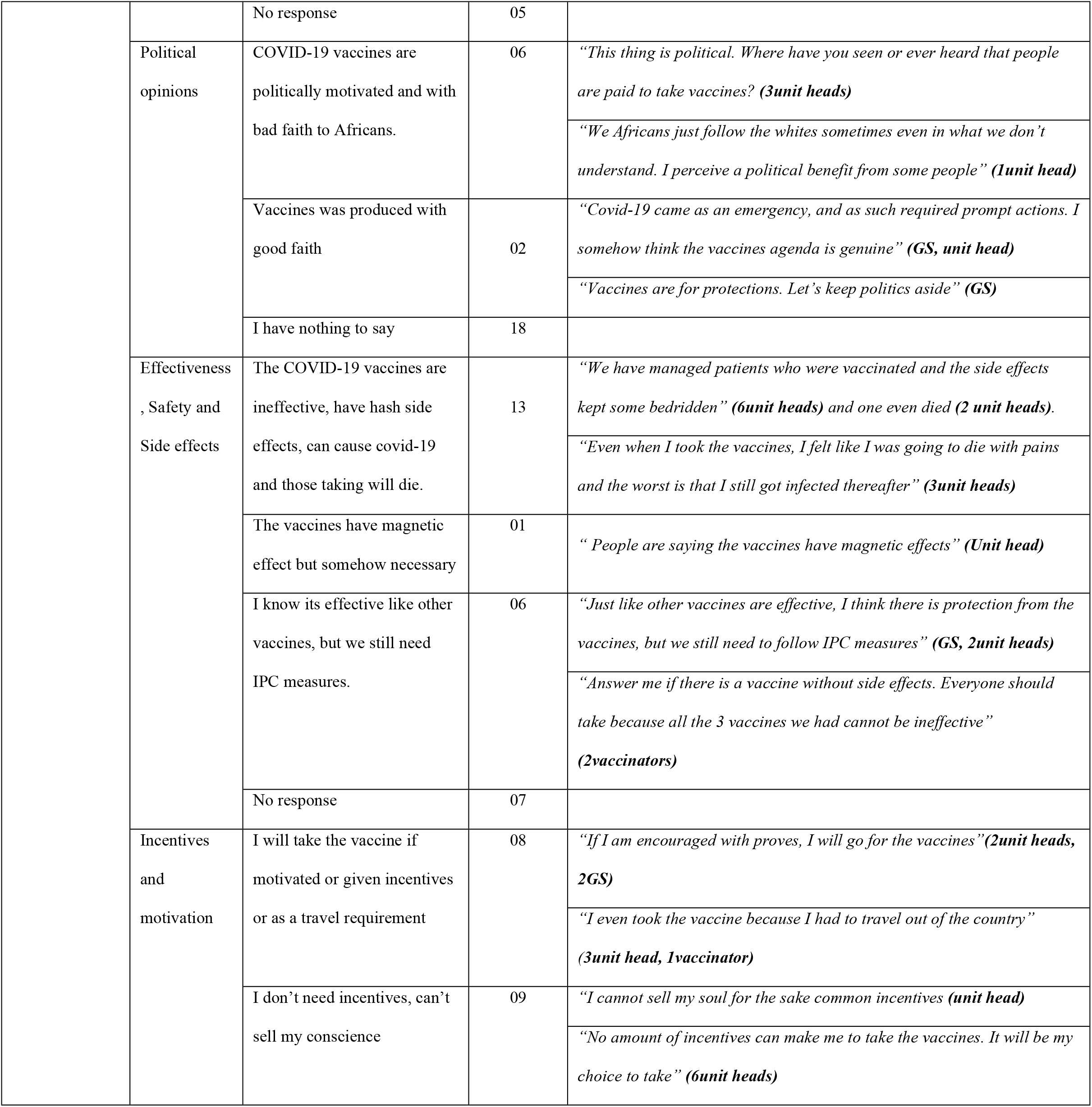
Nurse leaders’ perceptions regarding uptake of COVID-19 vaccines.

### Proportion of nurses and nurse leaders’ COVID-19 vaccination status in Fako Division

Up to 109 (55.3%) amongst the study population of nurses had not been vaccinated. Among the unvaccinated nurses, reasons given were: “we are not prepared” 40(20.3%), no psychological support 37(18.8%) and reluctant/not yet convinced to take the vaccines 32(16.2%) respectively. The majority of the nurses 40(45.4%), actually took the “Johnson & Johnson” vaccine followed by the Chinese “Sinopharm” 25(28.4%), with the least utilized vaccines being the “Pfizer” 23(26.2%)

In contrast, a majority 16 (61.54%) of the nurse leaders said *“I have been vaccinated against the COVID-19 disease with Johnson and Johnson (2 GS, 4unit heads, 2 vaccinators), Sinopharm (1vaccinator, 6unit heads) and AstraZeneca (vaccinator)”*. Most nurse leaders 09(34.62%) took the vaccines in 2022 as opposed to 2021 without being threatened or physically motivated 14(53.85%) to do so by anyone.

### Factors influencing COVID-19 vaccines uptake among nurses in Fako Division

Most nurses had the belief that the COVID-19 vaccines were dangerous and could cause death 120 (60.9). With social influence, most nurses said the government cannot influence them to take the vaccines (157, (79.7%). However, they reported that they would have taken the vaccines if they saw the need. 51(25.9%) of our respondents said they could be influenced to take the vaccines through the inspiration of their mentors, celebrity 36(18.3%) and politician 21(10.7%), while others 32(16.2%) said, none of these personalities can make them to change their minds (Tab 7).

**Table 4:** Factors Associated with COVID-19 Vaccines Uptake Among Nurses in Fako Division.

| Factors | Responses from the respondent | Frequency n (%) |
| --- | --- | --- |
| **Belief factor** | Dangerous / Cause Death | 120 (60.9) |
|  | Risky Vaccines | 22 (11.2) |
|  | Doubt Effectiveness | 55 (27.9) |
|  | **Total** | **197 (100.0)** |
| **Social Influence** | Government cannot influence me. I don't trust in what they say. | 157 (79.7) |
|  | Government can influence me to take. | 40 (20.3) |
|  | If others refuse the COVID-19 vaccines, I will still take. | 140 (71.1) |
|  | I will believe in these social figures to take the COVID-19 vaccines; |  |
|  | - Politician | 21 (10.7) |
|  | - Academic Mentor | 51 (25.9) |
|  | - Celebrity | 36 (18.3) |
|  | None of them. We have different immune systems | 32 (16.2) |
| **Knowledge and attitude** | Knowing about the vaccines can prompt me to take especially if it is good. | 82 (41.6) |
|  | Whether correct or incorrect knowledge about the vaccines, I cannot take. | 115 (58.4) |
|  | I know the benefits of being vaccinated. It's for immunization | 107 (54.3) |
|  | I cannot be certain | 90 (45.7) |
| **Religion Values** | My religious values and faith concerning the vaccines, limits me to take, though I can't be certain to explain. | 46 (23.4) |
|  | No religious values | 151 (76.6) |

### Factors influencing COVID-19 vaccines uptake among nurse leaders in Fako Division

The respondents reported different factors (belief factors, social influence and knowledge), that had negatively influenced them from taking the COVID-19 vaccines (Tab 10).

**Table 5:**
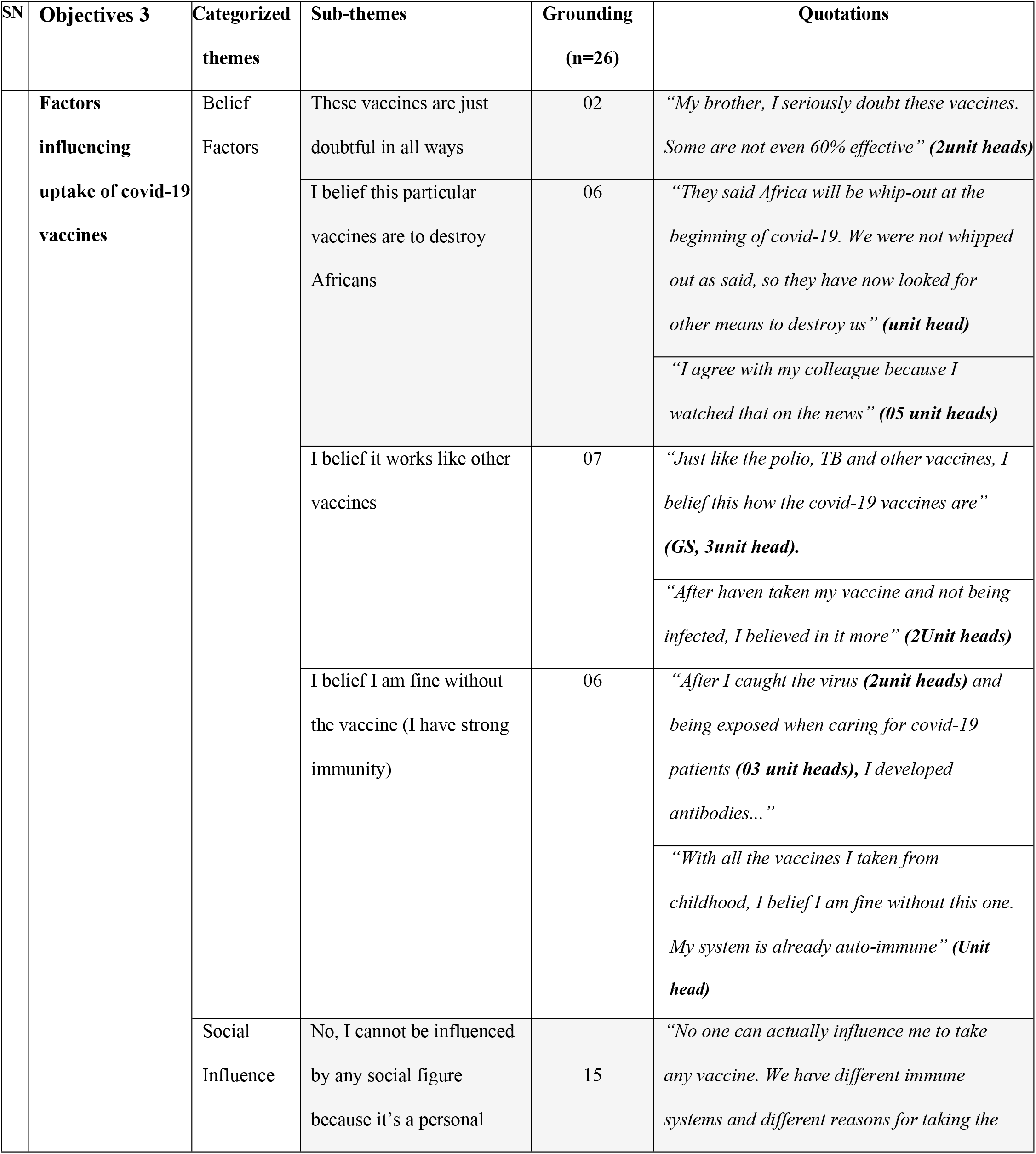
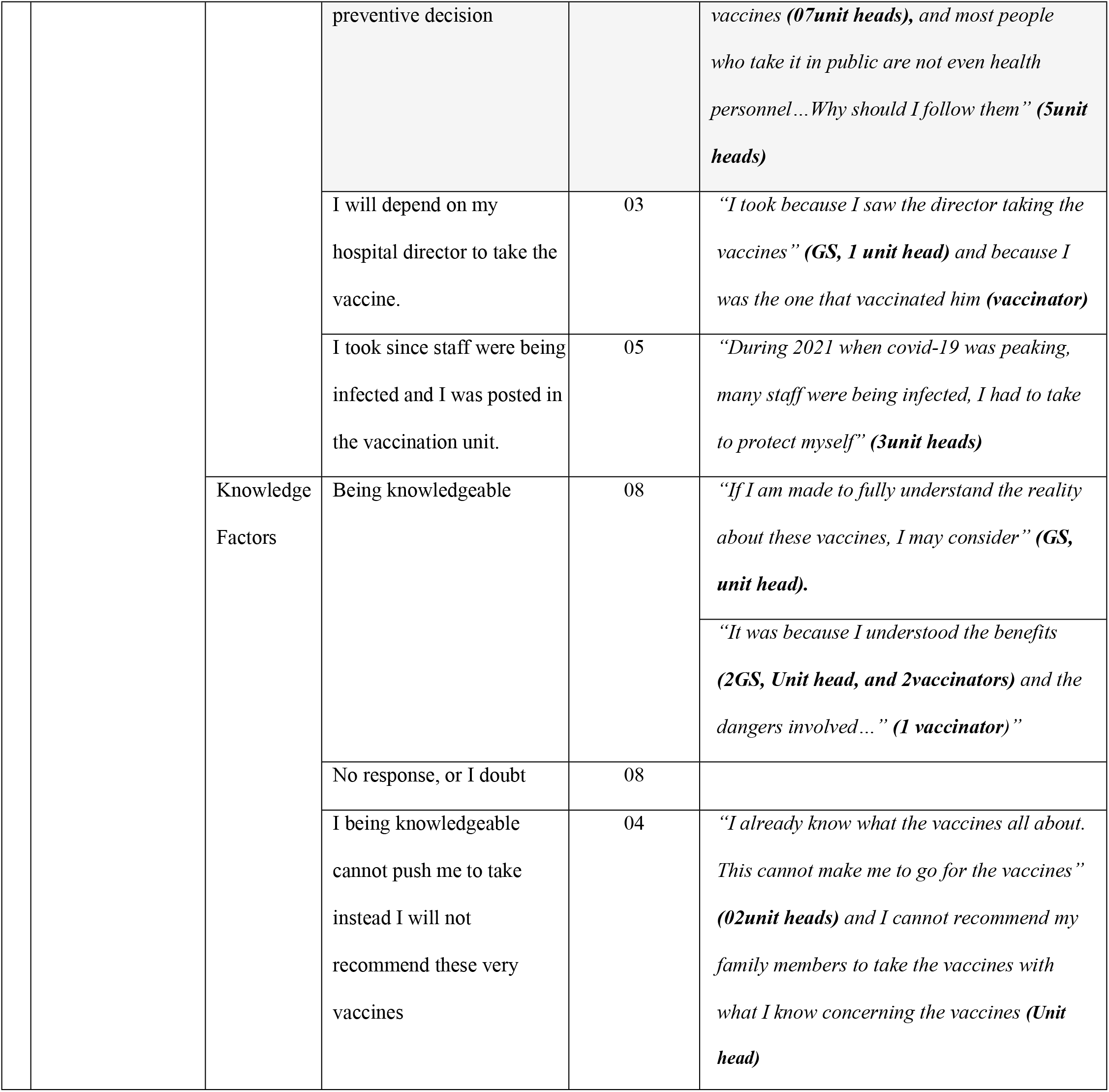
Factors influencing COVID-19 vaccines uptake among nurse leaders in Fako Division

## Discussion

### Nurses’ perceptions regarding the uptake of covid-19 vaccine

In this study, 133 (67.5%) nurses expressed positive perceptions about the vaccines; 74(37.5%) were confident to take the vaccines when exposed to infected cases, 25(16.8%) will take if they understood its benefits and only 15(07.6%) will take the vaccines if the disease proves severe on others. A few of them said they were overwhelmingly confident 19(05.6%) about taking the vaccines. In a similar study conducted in Philadelphia, United States (US) [17], 63.7% of employees reported that they planned and would want to receive a COVID-19 vaccine, 26.3% were unsure, and 10.0% did not plan to be vaccinated. In another study conducted in Saudi Arabia, it was observed that 50.5% were willing to have the COVID-19 vaccine with up to 49.7% intending to take the vaccine as soon as it became available in the country, while 50.3% indicated they would delay until the vaccine’s safety was confirmed [18]. Since the COVID-19 disease was new to every nation, its perceieved threat and severity in Cameroon, the USA and Saudi Arabia between 2020 to 2022 was devasting. Consequently, individuals were expected to consider the benefits of taking the vaccines rather than envisaging the risk(s) involved with vaccine uptake.

In contrast, a study conducted in the South West Region (SWR) of Cameroon revealed that the positivity rate of COVID-19 dropped from 17.7% in December 2021 to 4.1% in January 2022 [19]. This finding is in contrast with our results probably because of the declining transmission of COVID-19 in the SWR in 2022. Consequently, residents did not see the need to take the vaccine.

Even with nurses’ willingness for a COVID-19 vaccine, they had negative perceptions regarding its uptake. Majority reported; no trust in the ingredients & pharmaceutical companies 26(07.6%); vaccines could not be safe, ineffective and the presents of dangerous side effects 24(25.9%); and could cause COVID-19 infection 07(6.6%). This is similar to [20], where several nurses had negative perceptions regarding the uptake of a COVID-19 vaccine. Measure reasons were skepticism and side effects (36%); 20.0% preferred using few medications to prevent themselves; 19% preferred gaining natural immunity, 22.0% preferred to wait because vaccine is new, and 18.0% did not trust the health care system. Moreover, others indicated that they were afraid of needle pain during vaccination (7/38), percieved ineffectiveness of the vaccines (7/38=18.42%), perceived low severity (5/38=13.16%) and distrust of government sources (5/38=13.16%)[21]. Consequently, nurses were considered to have understood that no medication or vaccines exist without side effects. However, the preventive purpose had always been greater.

### Proportion of nurses vaccinated against COVID-19

In this study, only 88 (44.7%) of nurses in the quantitative phase had taken a COVID-19 vaccine. Similar to a report by the Ministry of Health (MoH) in Cameroon and WHO [19], only 1,812 doses of COVID-19 vaccines per 100,000 inhabitants, had been administered, representing 1.2% of the target population in Cameroon. The Ministry of Health (MoH) attributed the low vaccination rate to be associated with a small number of health staff administering vaccines, and the reluctance of the population to get vaccinated including HCWs. In the South West Region (SWR), where this study was conducted, the vaccination coverage remained low at 1.8% [19]. This low uptake of a COVID-19 vaccine by the population would have been ameliorated when nurses began to take the vaccines as frontline care givers and lead by example. In contrast to other studies, 32,903,329 people in South Africa (SA) had received 1 or more dose of a COVID-19 vaccine dose, giving the prevalence at 62.9% [22]. This sharp contrast could have possibly occurred due to the perceived benefits of being vaccinated and as a cue to action against the omicron BA.4 and BA.5 variant in April 2022, that increased the rate of infection in SA from 300 infections to 8.000 per week.

### Factors associated with COVID-19 vaccines uptake among nurses/nurse leaders

In this study, a majority of nurses and nurse leaders had the belief that the COVID-19 vaccines were dangerous and could result to killing someone 120 (60.9%). In a similar study among HCWs who refused vaccination, reported fear of side effects 67(93%), and the belief that the development time of the vaccines against COVID-19 was short, which could cause harm to people 69 (96.0%) [23]. Respondents in another study had the belief that the COVID-19 vaccines lack research and are risky (37.0%). Moreover, beliefs about rushed vaccine development, was the principal fact sorted and this alone could cause tremendous disaster to the population [24]. We realized that emergency vaccines were needed because of the insurgency of the COVID-19 disease, as such, every belief system would not have really been overemphasized as there was a great need for people to be safe through vaccination in every part of the world. There was no study within our search, published or unpublished, to contradict these findings.

Social influence was poorly reported. Up to 157 (79.7%) of our nurses could not be influenced by the government to take the vaccines. Findings from a similar study in Canada revealed mistrust towards governments and public health bodies (including mentors) on COVID-19 vaccines uptake [25; 26]. This could have been due to perceived political unwillingness to approve other treatment options like the Samuel Kleda’s COVID-19 complementary medicine in Cameroon. In contrast, Grumbach *et al*., [27] in their San Francisco study revealed up to 87.5% of HCWs who had trust in what the authorities were saying regarding the uptake of COVID-19 vaccine. The authorities in San Francisco could have revealed their COVID-19 infection status, toke the COVID-19 jab in public space and asssured the population with a strong will to respond to any eventuality if any one gets sick due to the vaccines. This was likely not the case in Cameroon.

Regarding knowledge factor, nurses with either correct or incorrect knowledge about COVID-19 vaccine (115 (58.4%), did not agree to take the vaccines. According to a similar study [28], Up to 78% of HCWs reported that ‘not knowing enough about the vaccine’ was a barrier to vaccination. Not enough information (73%) was cited as reason for vaccine refusal [29] and others reported a lack of understanding about the vaccine containing live or attenuated virus [26]. Not knowing about COVID-19 vaccines and its potency can greatly infuence the nurse not to take the vaccines. In cameroon, being knowledgeable or not as a nurse is not a pre-requisite to take a COVID-19 vaccine. This was because, nurses had possibly not understood the motive for the sudden pressure of all frontline care-givers in Cameroon to take a COVID-19 vaccine when other measures than vaccination proved to have saved more lives.

## Conclusion

Nurses perceived high relevance for the COVID-19 vaccines 133(67.51%) while a majority of the nurse leaders perceived that the COVID-19 vaccines were; ineffective with dangerous side effects; no trust in the ingredients and pharmaceutical industries; and the vaccines could have the possibility of causing COVID-19 infection. Even with the negative perceptions, most nurses and nurse leaders were still willing to take the vaccines once they understood its benefits and its side effects.

A significant proportion of nurses in Fako Division, had not yet been vaccinated against COVID-19. However, a majority of nurse leaders (unit heads, GS and COVID-19 vaccination unit coordinators) had been vaccinated with one or two of the currently available COVID-19 vaccines. Negative factors that influenced COVID-19 vaccines uptake by nurses and nurse leaders in Fako division, included the belief that the vaccines were dangerous and could cause death. There were also religious and social factors reported, which could be addressed if we must increase COVID-19 vaccination uptake and deal with the multi-faceted impact of COVID-19 infection in Fako Division.

## Data Availability

All data can be accessed on the software used for analysis

## Acknowledgement

I immensely appreciate my supervisors Prof. Palle John Ngunde, who doubles as the Head of Nursing Department and Dr. Binwi Florence Nkemayim for their corrections, guidance, words of inspiration and time allocated to the successful completion of this scientific thesis. My deepest and warmest thanks to all those who participated in this study at the Tiko District Hospital, Limbe and Buea Regional Hospitals and also the statisticians who diligently analyzed this work. Achidi EA is a Co-investigator of the central Africa Network for TB, HIV/AIDS and Malaria (CANIAM), a regional network of excellence grant from the European and Developing Countries Trials Partnership (EDICTP)

## Authors’ contribution

**Conceptualization** Niba Clinton Ambe

**Data curation:** Niba Clinton Ambe

**Formal analysis:** Niba Clinton Ambe, Achidi Eric Akum

**Investigation:** Niba Clinton Ambe, Achidi Eric Akum

**Methodology:** Niba Clinton Ambe, Nkemayim Florence Binwi, Palle John Ngunde, Achidi Eric Akum

**Software:** Niba Clinton Ambe, Achidi Eric Akum

**Supervision:** Nkemayim Florence Binwi, Palle John Ngunde

**Validation:** Niba Clinton Ambe, Nkemayim Florence Binwi, Palle John Ngunde, and Achidi Eric Akum

**Visualization:** Niba Clinton Ambe, Nkemayim Florence Binwi

**Writing – original draft:** Niba Clinton Ambe

**Writing – review & editing:** Niba Clinton Ambe, Nkemayim Florence Binwi, Palle John Ngunde and Achidi Eric Akum

## References

1. Morens DM, Breman JG, Calisher CH, Doherty PC, Hahn BH, Keusch GT, Kramer LD, LeDuc JW, Monath TP, Taubenberger JK. The origin of COVID-19 and why it matters. The American journal of tropical medicine and hygiene. 2020 Sep;103(3):955.

2. Alanagreh LA, Alzoughool F, Atoum M. The human coronavirus disease COVID-19: its origin, characteristics, and insights into potential drugs and its mechanisms. Pathogens. 2020 Apr 29;9(5):331.

3. WHO. 2022: Cameroon Emergency response; Health. Available at https://reports.unocha.org/en/country/cameroon/#cf-1D7RNQ8SRMtdbV8jbRMlIx [Accessed 15th March 2022].

4. Liang W, Liang H, Ou L, Chen B, Chen A, Li C, Li Y, Guan W, Sang L, Lu J, Xu Y. China Medical Treatment Expert Group for COVID-19. Development and validation of a clinical risk score to predict the occurrence of critical illness in hospitalized patients with COVID-19. JAMA Intern Med. 2020 Aug 1;180(8):1081–9.

5. Guo T, Fan Y, Chen M, Wu X, Zhang L, He T, Wang H, Wan J, Wang X, Lu Z. Cardiovascular implications of fatal outcomes of patients with coronavirus disease 2019 (COVID-19). JAMA cardiology. 2020 Jul 1;5(7):811–8.

6. Belete TM. Review on up-to-date status of candidate vaccines for COVID-19 disease. Infection and drug resistance. 2021;14:151.

7. Stanley P, Plotkin SA. History of vaccination. Proceedings of the National Academy of Sciences. 2014 Aug 26;111(34):12283–7.

8. Omid V. E, Miriam S. Johnson, Sara Ebling, Ole M A, Øyvind H, Asle H, Nora S, and Sverre UJ. Risk, Trust, and Flawed Assumptions. Vaccine Hesitancy During the COVID-19 Pandemic. Front. Public Health, 01 July 2021 | https://doi.org/10.3389/fpubh.2021.700213

9. Omer SB, Salmon DA, Orenstein WA, Dehart MP, Halsey N. Vaccine refusal, mandatory immunization, and the risks of vaccine-preventable diseases. New England Journal of Medicine. 2009 May 7;360(19):1981–8.

10. Lin Y, Hu Z, Zhao Q, Alias H, Danaee M, Wong LP. Understanding COVID-19 vaccine demand and hesitancy: A nationwide online survey in China. PLoS neglected tropical diseases. 2020 Dec 17;14(12):e0008961.

11. Dror AA, Eisenbach N, Taiber S, Morozov NG, Mizrachi M, Zigron A, Srouji S, Sela E. Vaccine hesitancy: the next challenge in the fight against COVID-19. European journal of epidemiology. 2020 Aug;35(8):775–9. https://doi.org/10.1007/s10654-020-00671-y. Available from https://link.springer.com/article/10.1007/s10654-020-00671-y#citeas [accessed 17 February 2022].

12. Hagan K, Forman R, Mossialos E, Ndebele P, Hyder AA, Nasir K. COVID-19 vaccine mandate for healthcare workers in the United States: a social justice policy. Expert review of vaccines. 2022 Jan 2;21(1):37–45.

13. Nzaji MK, Ngombe LK, Mwamba GN, Ndala DB, Miema JM, Lungoyo CL, Mwimba BL, Bene AC, Musenga EM. Acceptability of vaccination against COVID-19 among healthcare workers in the Democratic Republic of the Congo. Pragmatic and observational research. 2020;11:103.

14. Dinga JN, Sinda LK, Titanji VP. Assessment of vaccine hesitancy to a COVID-19 vaccine in Cameroonian adults and its global implication. Vaccines. 2021 Feb 19;9(2):175. https://doi.org/10.3390/vaccines9020175 Ngasa NC, Ngasa SN, Tchouda LA, Tanisso E, Abanda C, Dingana TN. Spirituality and other factors associated with COVID-19 Vaccine Acceptance amongst Healthcare Workers in Cameroon. Retrieved on 15 February 2022 from https://www.researchsquare.com/article/rs-712354/v1

15. Israel GD. University of Florida IFAS Extension. Determining the sample size. 2009. Available from https://www.tarleton.edu/academicassessment/documents/samplesize.pdf [Accessed 12th May 2022].

16. Kuter BJ, Browne S, Momplaisir FM, Feemster KA, Shen AK, Green-McKenzie J, Faig W, Offit PA. Dinga JN, Sinda LK, Titanji VPPerspectives on the receipt of a COVID-19 vaccine: A survey of employees in two large hospitals in Philadelphia. Vaccine. 2021 Mar 19;39(12):1693–700.

17. Qattan AMN, Alshareef N, Alsharqil O, Al Rahahleh N, Chirwa GC & Al-Hanawil MK. 2021. Dinga JN, Sinda LK, Titanji VPAcceptability of a COVID-19 Vaccine Among Healthcare Workers in the Kingdom of Saudi Arabia. Front. Med., 01 March 2021 | https://doi.org/10.3389/fmed.2021.644300

18. WHO. Camerooon: North-West and South-West Health Cluster COVID-19 Epidemiological Bulletin. 2021. Retrieved on 14 March 2022 from https://www.humanitarianresponse.info/sites/www.humanitarianresponse.info/files/documents/files/nwsw_covid_bulletin_september.pdf

19. Jasuja GK, Meterko M, Bradshaw LD, Carbonaro R, Clayman ML, LoBrutto L, Miano D, Maguire EM, Midboe AM, Asch SM, Gifford AL. Attitudes and intentions of US veterans regarding COVID-19 vaccination. JAMA network open. 2021 Nov 1;4(11):e2132548..

20. Yaqub O, Castle-Clarke S, Sevdalis N, Chataway J. Attitudes to vaccination: a critical review. Soc Sci Med. 1982;2014(112):1–11.

21. Ministry of Health, Republic of South Africa. Availabel at https://sacoronavirus.co.za/ [accessed 17th March 2022].

22. Papagiannis D, Malli F, Raptis DG, Papathanasiou IV, Fradelos EC, Daniil Z, Rachiotis G, Gourgoulianis KI. Assessment of knowledge, attitudes, and practices towards new coronavirus (SARS-CoV-2) of health care professionals in Greece before the outbreak period. International journal of environmental research and public health. 2020 Jul;17(14):4925.

23. Tulloch JS, Lawrenson K, Gordon AL, Ghebrehewet S, Ashton M, Peddie S, Parvulescu P. COVID-19 vaccine hesitancy in care home staff: a survey of Liverpool care homes. MedRxiv. 2021 Jan 1.

24. Fouogue JT, Noubom M, Kenfack B, Dongmo NT, Tabeu M, Megozeu L, Alima JM, Fogang YF, Nyam LC, Fouelifack FY, Fouedjio JH. Poor knowledge of COVID-19 and unfavourable perception of the response to the pandemic by healthcare workers at the Bafoussam Regional Hospital (West Region-Cameroon). The Pan African Medical Journal. 2020;37(Suppl 1).

25. SafeCare-BC. Briefing Note: COVID-19 Vaccine Survey. SafeCare-BC 2020.

26. Grumbach K, Judson T, Desai M, Jain V, Lindan C, Doernberg SB, Holubar M. Association of race/ethnicity with likeliness of COVID-19 vaccine uptake among health workers and the general population in the San Francisco Bay Area. JAMA internal medicine. 2021 Jul 1;181(7):1008–11.

27. Kuter BJ, Browne S, Momplaisir FM, Feemster KA, Shen AK, Green-McKenzie J, Faig W, Offit PA. Perspectives on the receipt of a COVID-19 vaccine: A survey of employees in two large hospitals in Philadelphia. Vaccine. 2021 Mar 19;39(12):1693–700.

28. Ledda C, Costantino C, Cuccia M, Maltezou HC, Rapisarda V. Attitudes of Healthcare Personnel towards Vaccinations before and during the COVID-19 Pandemic. International Journal of Environmental Research and Public Health. 2021 Mar 8;18(5):2703.

